# Intra-county modeling of COVID-19 infection with human mobility: assessing spatial heterogeneity with business traffic, age and race

**DOI:** 10.1101/2020.10.04.20206763

**Authors:** Xiao Hou, Song Gao, Qin Li, Yuhao Kang, Nan Chen, Kaiping Chen, Jinmeng Rao, Jordan S. Ellenberg, Jonathan A. Patz

## Abstract

The novel coronavirus disease (COVID-19) pandemic is a global threat presenting health, economic and social challenges that continue to escalate. Meta-population epidemic modeling studies in the susceptible-exposed-infectious-removed (SEIR) style have played important roles in informing public health and shaping policy making to mitigate the spread of COVID-19. These models typically rely on a key assumption on the homogeneity of the population. This assumption certainly cannot be expected to hold true in real situations; various geographic, socioeconomic and cultural environments affect the behaviors that drive the spread of COVID-19 in different communities. What’s more, variation of intra-county environments creates spatial heterogeneity of transmission in different sub-regions. To address this issue, we develop a new human mobility flow-augmented stochastic SEIR-style epidemic modeling framework with the ability to distinguish different regions and their corresponding behavior. This new modeling framework is then combined with data assimilation and machine learning techniques to reconstruct the historical growth trajectories of COVID-19 confirmed cases in two counties in Wisconsin. The associations between the spread of COVID-19 and human mobility, business foot-traffic, race & ethnicity, and age-group are then investigated. The results reveal that in a college town (Dane County) the most important heterogeneity is spatial, while in a large city area (Milwaukee County) ethnic heterogeneity becomes more apparent. Scenario studies further indicate a strong response of the spread rate on various reopening policies, which suggests that policymakers may need to take these heterogeneities into account very carefully when designing policies for mitigating the spread of COVID-19 and reopening.

## 1 Introduction

The novel coronavirus disease (COVID-19) pandemic is a global threat presenting health, economic and social challenges that continue to escalate. As of September 30, 2020, the CDC had reported 7,168,077 total confirmed cases and 205,372 total deaths in the U.S.^1^, and community transmission of COVID-19 remains widespread in many parts of the world. In the absence of vaccines or pharmacologic agents to reduce the transmission of the severe acute respiratory syndrome coronavirus-2 (SARS-CoV-2) that causes COVID-19, it is essential to understand the effects of non-pharmacological epidemic control and intervention measures. These include, but are not limited to, social (physical) distancing, travel restrictions, closures of schools and nonessential business services, mandated face coverings, testing, isolation, contact tracing and timely quarantine on delaying the COVID-19 spread, all of which have been intensely investigated^2–8^. In the end, the same combination of interventions will have different effects on the overall progress of the epidemic in different contexts, and the dependence of outcomes on local conditions is typically complex. It is thus essential to develop good *models* which can make principled predictions about the effect of non-pharmacological interventions (NPIs) on the spread of COVID-19 under various combinations of local circumstances. For example, during the early outbreak in China, scholars developed a model using observed data about human mobility and COVID-19 infection and found that early detection and isolation of confirmed cases could prevent more infections than travel restrictions and contact reductions, but that combined NPIs achieved the strongest and most rapid effect^4^. Another study using laboratory-confirmed cases demonstrated the effects of various types of public health NPIs (such as centralized quarantine and isolation) on reducing the transmission of the SARS-CoV-2) in Wuhan, China^2^. In Italy, scholars reconstructed the COVID-19 spatial spread dynamics and investigated the effects of population-wide interventions^9,10^.

The large body of modeling work on COVID can be grouped into four broad categories: mechanistic models^2,4,6,8–13^, agent-based models^14,15^, generalized linear regression models^16,17^, and machine learning models^18,19^. Both mechanistic and agent-based models typically rely on susceptible-infectious-removed (SIR) or susceptible-exposed-infectious-removed (SEIR) compartmental-style models in epidemiology, which separates the population into four compartments (susceptible, exposed, infectious, recovered or removed) and models the dynamics of transitions between the compartments. In the U.S., the Institute for Health Metrics and Evaluation (IHME) at the University of Washington also employed a SEIR-style framework to project hospital bed-days, ICU-days, ventilator days and deaths, and to model possible trajectories of SARS-CoV-2 infections and the impact of NPIs at the state level in the U.S^11,12^. Regarding the time-varying state-specific control measures, a temporal extended susceptible-antibody-infectious-removed (eSAIR) model was developed for the projection of county-level COVID-19 prevalence^20^. Under the agent-based modeling framework, a recent study modeled the impact of testing, contact tracing and household quarantine on the second waves of COVID-19 infection in the Boston metropolitan area^14^.

However, a straightforward SEIR analysis, useful as it may be in revealing and informing public health and shaping policy making, relies on a key assumption of homogeneity of the population. Each compartment is treated as an aggregate of indistinguishable individuals. This is not realistic. A recent study has shown evidence of spatial heterogeneity in COVID-19 transmissions within cities of Seattle and Washington, DC^21^. Socioeconomic and cultural environments, which differ within small geographic communities, affect the behaviors that drive the spread of COVID-19. What’s more, geographic and transportation factors create spatial heterogeneity of infection spread in different sub-regions. Accurate and fine-grained modeling requires a more refined classification of the population.

Examining the spatial heterogeneity of infection spread is particularly important for modeling COVID-19 due to its complex dependence on social factors. COVID-19 presents a highly difficult policy issue because people need to weigh various trade-offs when they make health behavior decisions, trade-offs which may involve political ideologies, socioeconomic status and lifestyles^22^. Extensive research from social science has demonstrated that people’s assessments of these trade-offs and their resulting health and risk behavior are affected by a variety of factors, such as gender, race, political ideology, economic status, culture, and religion^23–27^. Despite a scientific consensus on the effectiveness of social distancing, precautionary actions like facial masking and social distancing are not universally adhered to,^28^, and adherence to these measures is by no means uniform across populations. Thus, modeling the spread of COVID-19 requires scholars to attend to the heterogeneity in people from different backgrounds, with different value systems, and in different locales. The place (i.e., the type of neighborhoods and communities) people live in is a reflection of race, socioeconomic status, and age^29,30^. Segregation along these lines is not just an aggregate of individual choices, but a consequence of public polices^31^. Emerging evidence has shown disparities in COVID-19 outcomes between social groups. For instance, scholars found that income, along with race and other socioeconomic factors, has become a major predictor of COVID-19 infections and deaths^32,33^. A team of international scholars has addressed the importance of using social and behavioral sciences to support COVID-19 pandemic responses^34^, such as the study of social inequalities and political polarization in response to social distancing and stay-at-home mandates^35–37^.

In this paper, we aim to capture the effect of social and geographic heterogeneity within a population, and to reveal multiple aspects of the influences from different cultural environments, by building models with finer spatial resolution within a small geographic region (in this case, a single county.) To this end, we have developed a new human mobility flow-augmented stochastic SEIR-style epidemic modeling framework that is able to distinguish different regions and their corresponding behavior. This new modeling framework is then combined with data assimilation and machine learning to reconstruct the historical growth trajectories of COVID-19 confirmed cases. We then investigate the associations between the spread of COVID-19 and human mobility, business foot-traffic, race, and age group. Finally, we perform scenario studies in order to understand the social and policy implications as well as to facilitate the development of guidelines to inform future policy-making in public health crisis. The two geographic regions we consider are Dane County, WI and Milwaukee County, WI, which contain the two largest cities in Wisconsin — respectively, Madison (a college town and home of the state government) and Milwaukee (a large city subject to severe racial and ethnic segregation, by some measures the most extreme in the United States^38,39^.)

## 2 Data

The official testing results of COVID-19 confirmed cases between March 11, 2020 and August 12, 2020 were obtained from the local COVID-19 Dashboards, created by the Public Health Offices of City of Madison & Dane County and Milwaukee County^40,41^. The census-tract level geographic boundaries with demographics and socioeconomic attributes were obtained from the U.S. Census Bureau^42,43^. We collected over 3.6 million points of interest (POIs) with aggregated and de-identified human travel patterns in the U.S. from SafeGraph, which data was further spatially filtered by our study area. SafeGraph’s data is demographically distributed in a way that matches the proportion of people in various population subgroups in the United States Census populations^1^. These mobile location data consist of “pings” identifying the coordinates of a smartphone at a moment in time. To enhance privacy, SafeGraph excludes census block group (CBG) information if fewer than five devices visited a place in a month from a given CBG. For each POI, the records of aggregated visitor patterns record the number of unique visitors and the number of total visits to each venue during a specified time window (i.e., hourly, weekly, and monthly); this allows us to estimate the foot-traffic of each venue and the origin-to-destination (O-D) spatial interaction flow patterns during the the study period^44^. We further aggregate the O-D flow matrices to the census-tract level to match the COVID-19 testing data.

## 3 Mathematical model and configuration

In this research we develop a new human mobility flow-augmented stochastic SEIR model to simulate and analyze the deep connection between the fast spread of COVID-19 and individual mobility, social activities, government orders and population distribution, using Milwaukee County and Dane County, the two most populated counties in the state of Wisconsin as examples. The classical susceptible-exposed-infectious-removed (SEIR) model divides the population into four compartments, *S* (susceptible), *E* (latent), *I* (reported infections) and *R* (removed from *I*) and uses an ordinary differential equation (ODE) system to describe the flow between the compartments. The model assumes the homogeneity within the population, but this assumption is typically not valid. Our new approach is to investigate the compartment model at a finer scale, to reveal the differences and connections between sub-communities within one county, and to recover the dynamics of the population for different sub-regions.

In our new approach, human mobility flow data is incorporated into machine learning clustering algorithms first to divide a county into several regions that present diverse social, cultural, and economical background. We then assume the homogeneity within each smaller region. Human mobility flow-augmented stochastic SEIR models are then built for each region respectively. The models are coupled together through mobility data that adds strong nonlinear interactions between regions into the model. Figure S1 demonstrates the flowchart of this model.

It is worthwhile to highlight the following key features in this new human mobility flow-augmented stochastic SEIR model. We use a simple but effective stochastic (Ornstein-Uhlenbeck) process to describe the dynamical evolution of key parameters in the model, e.g. the transmission rate. This it to contrast the use of a constant transmission rate as is done in the classical SEIR model, and a function of time without any structure as is done in most recent works^45^. Our approach not only reflects the fact that the parameters are not fixed constants, but rather adjust the values according to the changes of the environments, but also permits a cheap but effective online learning process of reconstruction. This reconstruction procedure will be discussed in section of parameter setting.

We now discuss the clustering process using human mobility data, and the mathematical model respectively.

### 3.1 Origin-to-Destination (O-D)-flow based spatial clusters

Each county to be studied will be divided into multiple smaller regions. We use the Walktrap network-based community detection method^46^ that utilizes short random walks to detect communities in a large graph. The travel O-D flow data of the first week of March (right before the widespread of COVID-19 in U.S.) is used as the graph weights for clustering. Denote *N* the number of census tracts (the smallest area unit) in the whole area. The algorithm starts with an initial division of the whole area into *N* regions, each containing only one census tract, and iterates till the best clustering is found. During each iteration, the distances between each pair of adjacent vertices are computed, upon which, by minimizing the mean of the squared distances between each vertex and its region, the distance between regions are calculated. The algorithm merges the adjacent regions so that upon merging, the smallest mean-squared distance is achieved. After *N* − 1 iterations, we acquire *N* different partitions and the one with smallest modularity is chosen as the product of the algorithm. The distance between vertex *i* and vertex *j* used in this algorithm is defined as

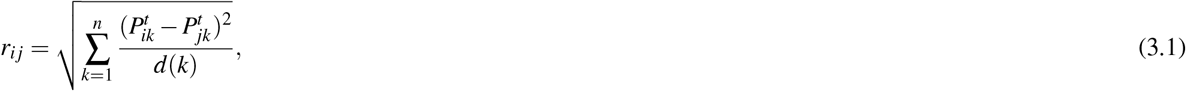

where 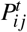 is the probability of a random walk to go from *i* to *j* in *t* steps and the degree *d*(*i*) is the number of neighbors of vertex *i*, including itself, and *t* is a user-chosen parameter. The Walktrap method utilizes the property that random walk tends to get “trapped” in more connected areas.

### 3.2 A new human mobility flow-augmented stochastic SEIR model

We denote different regions using sub-indices *i*, the new human mobility flow-augmented stochastic SEIR model on each region writes as:

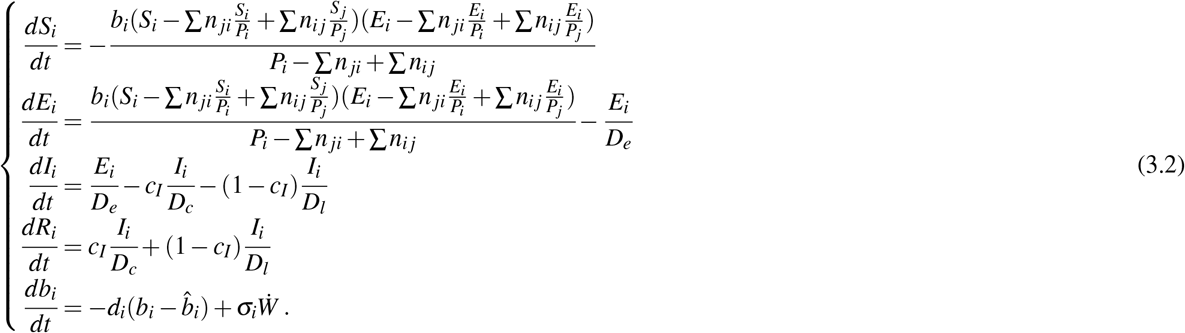

It is a coupled ODE-SDE (stochastic differential equation) system. *P*_*i*_ = *S*_*i*_ + *E*_*i*_ + *R*_*i*_ denotes the free population that can move between regions. We assume the reported cases are quarantined, and thus removed from *P*_*i*_. The parameters used in the model^15^ are explained below:

- *n*_*i j*_: the daily traffic flow from region *j* to region *i*.
- *D*_*e*_: the duration of latent period. We set *D*_*e*_ = 5.
- *D*_*c*_: the duration of infection for critical cases. We set *D*_*c*_ = 10.
- *D*_*l*_: the duration of infection for non-critical cases. We set *D*_*l*_ = 6
- *c*_*I*_: the proportion of critical cases among all reported cases. We assume *c*_*I*_ = 0.1.
- *b*_*i*_: the transmission rate in region *i*
- 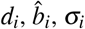: parameters in stochastic process for *b*_*i*_.

We use the following initial data (*t* = 0 at the start of the study period):

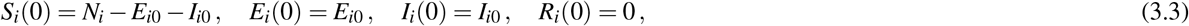

where the unit for *t* is one day and *N*_*i*_ is the total population of region *i*.

The effective reproduction number is one of the key quantities that reflects the speed of the spreading of the disease. It measures, on average, how many new cases are directly generated by a single infection. In our model, it can be computed by:

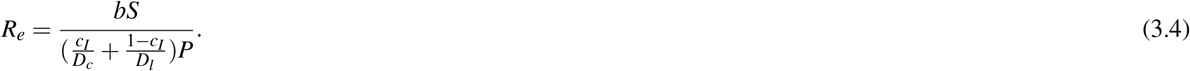

We note that the transmission rate *b* and ratio of susceptible *S*/*P* are different for each region, and thus each region has its own *R*_*e*_.

The feature that differentiates our approach from the classical SEIR model is our incorporation of population heterogeneity. This feature enables our new human mobility flow-augmented stochastic SEIR model to simulate and analyze the important differentiations between multiple sub-communities within one county, and the connection between spread of COVID-19 and local mobility and social activities within and among these subcommunities.

To be more specific: a traditional SEIR model divides the population into four compartments and uses an ordinary differential equation (ODE) system to describe the flow between the compartments. This treats the individuals within each compartment as indistinguishable, and thus misses important distinctions which are relevant to epidemic dynamics. By contrast, our model begins by using a machine learning algorithm to partition the county into clusters, using the observed human mobility flow data. Once these subcommunities have been generated, we build a local SEIR model for each region, which takes into account region-specific social, cultural, and economic factors; finally, we couple together all these local SEIR models using the data on intra-county mobility flow. The use of multiple non-identical subcommunities has a significant impact of the overall SEIR dynamics and its predictability.

### 3.3 An efficient data assimilation strategy for reconstructing infection trajectory

The application of data assimilation for the reconstruction of the remaining parameters in the model is another desirable feature of our approach. In particular, we apply the ensemble Kalman filter method^47^ to estimate the hidden parameters in the model, and to recover the trajectory of the spread of the disease. Such a model calibration procedure is completely data-driven, with no ad-hoc parameter tuning or adjustment. It is shown that the model with parameters estimated in this way can almost perfectly recover the observed time series data, which justifies the application of the data assimilation approach utilized here.

#### 3.3.1 Ensemble Kalman Filter

To estimate parameters and state variables, we use the ensemble Kalman Filter^47–49^. It was derived from the classical Kalman Filter for the application in atmospheric science, with the analytical covariance matrix replaced by its ensemble version, eliminating the computation of the Kalman gain matrix and the Riccati Equation that are typically expensive when problems are in high dimensional space. In each iteration, it combines the data with prior information to generate a posterior distribution of the state variables and parameters. The idea is to sample a fixed number of particles on the state variable space according to the initial distribution, and move these particles around at every time step, to obtain the adjusted distribution. Let *u* be the augmented vector of state variables and parameters for each region:

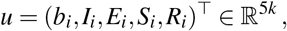

where *k* is the number of regions we get from clustering. The goal is to derive a distribution density function of *u*. At the beginning of each iteration, let *P*_*n*−1|*n*−1_ be the probability density to start with. Then we perform an evolution step to get the probability density *P*_*n*|*n*−1_ and a further analysis step to obtain *P*_*n*|*n*_. This serves as the initial probability density for the next iteration.

Let 𝒢_*m,n*_(*u*) be the solution to the model (3.2) that runs from time *t*_*m*_ to time *t*_*n*_ with *u* being the initial condition. Since the available data is the cumulative confirmation cases, we set 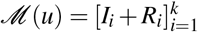 as our measuring operator, mathematically it is:

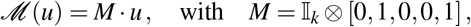

where 𝕀_*k*_ is an identity matrix of size *k*, and *M* maps a 5*k* dimensional vector to a *k* dimensional vector.

We assume that the infection data has a Gaussian noise from the true measuring operator:

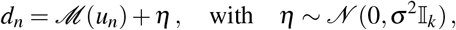

and thus the likelihood function is:

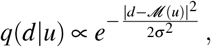

where ∝ is the proportional sign. The evolution step and analysis step in each iteration can be then calculated:

− Evolution: *P*_*n*|*n*−1_(*u*) = *δ* (*u*−𝒢_*n*−1,*n*_(*u*′))*P*_*n*−1|*n*−1_(*u*′),
− Analysis: *P*_*n*|*n*_(*u*) ∝ *P*_*n*|*n*−1_(*u*)*q*(*d*|*u*),

We need to further normalize to have ∫*P*_*n*|*n*_(*u*)*u* = 1.

Suppose we sample *N* particles, and we denote the *j*-th particle after the evolution step and analysis step at time *t*_*n*_ by 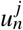 and 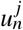 respectively. In summary, the algorithm iteratively applies to the following evolution and analysis steps:

− Evolution:

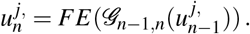

where *FE* is the Forward-Euler discretization applied to the model (3.2).

− Analysis:

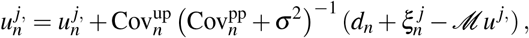

where 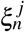is a *k*-length vector with each entry i.i.d. drawn from 𝒩 (0, *σ*^2^). *d*_*n*_ is the *k*-length vector collecting the reported infected data on *k* regions on day *n*. To compute the covariance matrices, we set:

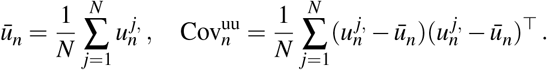

Then

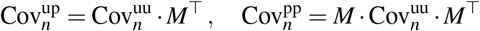

are matrices of size 5*k* × *k* and *k* × *k* respectively.

#### 3.3.2 Online learning of the transmission rate using an effective stochastic parameterization approach

One fundamental difference between our model and the classical SEIR model is that we embed stochasticity in the to-be-reconstructed parameters, the transmission rate, in particular. The last equation in (3.2) exploits a possibility of including the time variation in *b*. This is crucial in describing the transient behavior of the entire system. Since the exact governing law of the transmission rate is unknown and it is not a directly observed model variable, there is no need to describe the transmission rate in an exact fashion. Thus, an effective way of modeling the transmission rate is to adopt a simple stochastic parameterized form, which allows a cheap online learning process. To incorporate the stochastic parameterization in a simple and effect fashion, an Ornstein–Uhlenbeck (OU) process is adopted, which involves a Gaussian process as the prior ansatz of the transmission rate. The exact time evolution of the transmission rate, however, can be arbitrary. The parameters in the stochastic transmission rate equation is determined systematically using an expectation-maximization (EM) algorithm^50^, which is completely data-driven with no ad-hoc tuning or adjustment.

One starts from an arbitrary initial guess of the three parameters *d*_*i*_, *σ*_*i*_, and 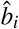 in the stochastic parameterized equation (the last equation in (3.2). In each EM iteration, one applies Ensemble Kalman Filter, resulting an estimated time series for each state variable and the transmission *b*_*i*_, and these parameters of OU process are then updated to fit the time series of *b*_*i*_ by matching the mean, variance, and decorrelation time. The fitting strategy can be viewed as the maximization process, and at the end of iteration, the optimal configuration of the three parameters, up to a preset threshold error, is found. This configuration is finally used in the last stage of Ensemble Kalman Filter to acquire the estimation of state variables and transmission rates used in our study.

## 4 Results

### 4.1 Origin-to-destination flow based spatial clusters

The traditional SEIR model assumes homogeneity over its population. This assumption, however, is not valid in either of the two counties studied here. To have a finer scale study, we divide both counties into several smaller intra-county regions using the Walktrap network-based community detection method^46^. The travel O-D flow data of the first week of March (just before the initial widespread outbreak of COVID-19 in the U.S.) is used to generate the graph weights for clustering.

The spatial clustering results for the two counties under investigation are shown in Figure 1b and Figure 1d. Dane County is partitioned into 7 regions (the two white areas are lakes, Mendota and Monona). Each region has its unique cultural features. In this respect it is worth specificially singling out Region 7, the area containing downtown Madison and the University of Wisconsin-Madison campus. As one might expect, Region 7 has significantly higher population density than the others. Region 3 is the residential region adjacent to Region 7 and it also has relatively high population density. The data is seen in Table 1. Milwaukee County is partitioned into 6 regions. While the population density in each region is relatively uniform, the region is heavily segregated by race and ethnicity. The population density and the distribution of different race and ethnic backgrounds is listed in Table 2. Region 3 has a predominantly Black population, while Region 4 has a high concentration of Hispanic residents.

**Table 1.**
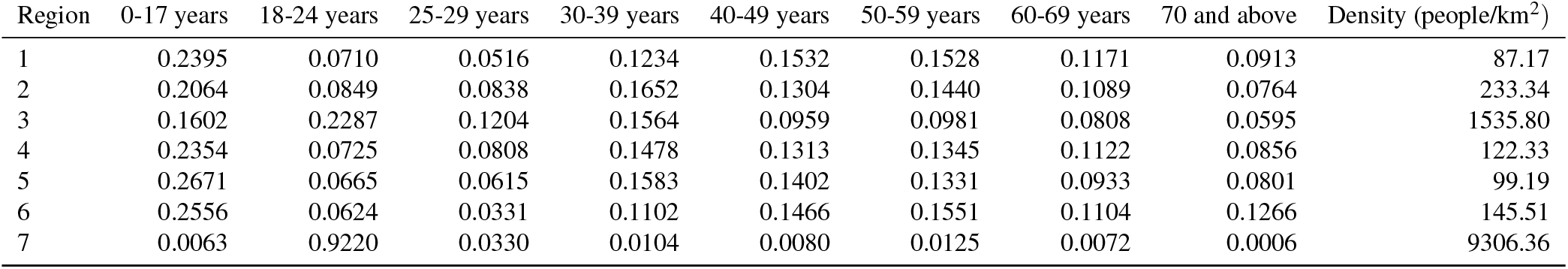
Population density and ratio of different age & ethnicity groups in Dane County.

| Region | 0-17 years | 18-24 years | 25-29 years | 30-39 years | 40-49 years | 50-59 years | 60-69 years | 70 and above | Density (people/km<sup>2</sup>) |
| --- | --- | --- | --- | --- | --- | --- | --- | --- | --- |
| 1 | 0.2395 | 0.0710 | 0.0516 | 0.1234 | 0.1532 | 0.1528 | 0.1171 | 0.0913 | 87.17 |
| 2 | 0.2064 | 0.0849 | 0.0838 | 0.1652 | 0.1304 | 0.1440 | 0.1089 | 0.0764 | 233.34 |
| 3 | 0.1602 | 0.2287 | 0.1204 | 0.1564 | 0.0959 | 0.0981 | 0.0808 | 0.0595 | 1535.80 |
| 4 | 0.2354 | 0.0725 | 0.0808 | 0.1478 | 0.1313 | 0.1345 | 0.1122 | 0.0856 | 122.33 |
| 5 | 0.2671 | 0.0665 | 0.0615 | 0.1583 | 0.1402 | 0.1331 | 0.0933 | 0.0801 | 99.19 |
| 6 | 0.2556 | 0.0624 | 0.0331 | 0.1102 | 0.1466 | 0.1551 | 0.1104 | 0.1266 | 145.51 |
| 7 | 0.0063 | 0.9220 | 0.0330 | 0.0104 | 0.0080 | 0.0125 | 0.0072 | 0.0006 | 9306.36 |

**Table 2.** Population density and ratio of different race & ethnicity in Milwaukee County.

| Region | White | Black | Native | Asian | Hawaii | Other | Hispanic | Density (people/km<sup>2</sup>) |
| --- | --- | --- | --- | --- | --- | --- | --- | --- |
| 1 | 0.75514 | 0.14589 | 0.00458 | 0.03943 | 0.00030 | 0.02326 | 0.06822 | 1623.00 |
| 2 | 0.87516 | 0.03936 | 0.00603 | 0.03445 | 0.00010 | 0.01736 | 0.11594 | 1029.98 |
| 3 | 0.17563 | 0.71883 | 0.00280 | 0.05269 | 0.00009 | 0.01718 | 0.04318 | 2032.80 |
| 4 | 0.66486 | 0.06772 | 0.00948 | 0.02575 | 0.00039 | 0.19042 | 0.48219 | 2473.66 |
| 5 | 0.81255 | 0.09696 | 0.00731 | 0.03292 | 0.00014 | 0.01629 | 0.06921 | 1719.89 |
| 6 | 0.89085 | 0.02476 | 0.00922 | 0.03776 | 0.00038 | 0.01056 | 0.09594 | 738.84 |

**Figure 1.**
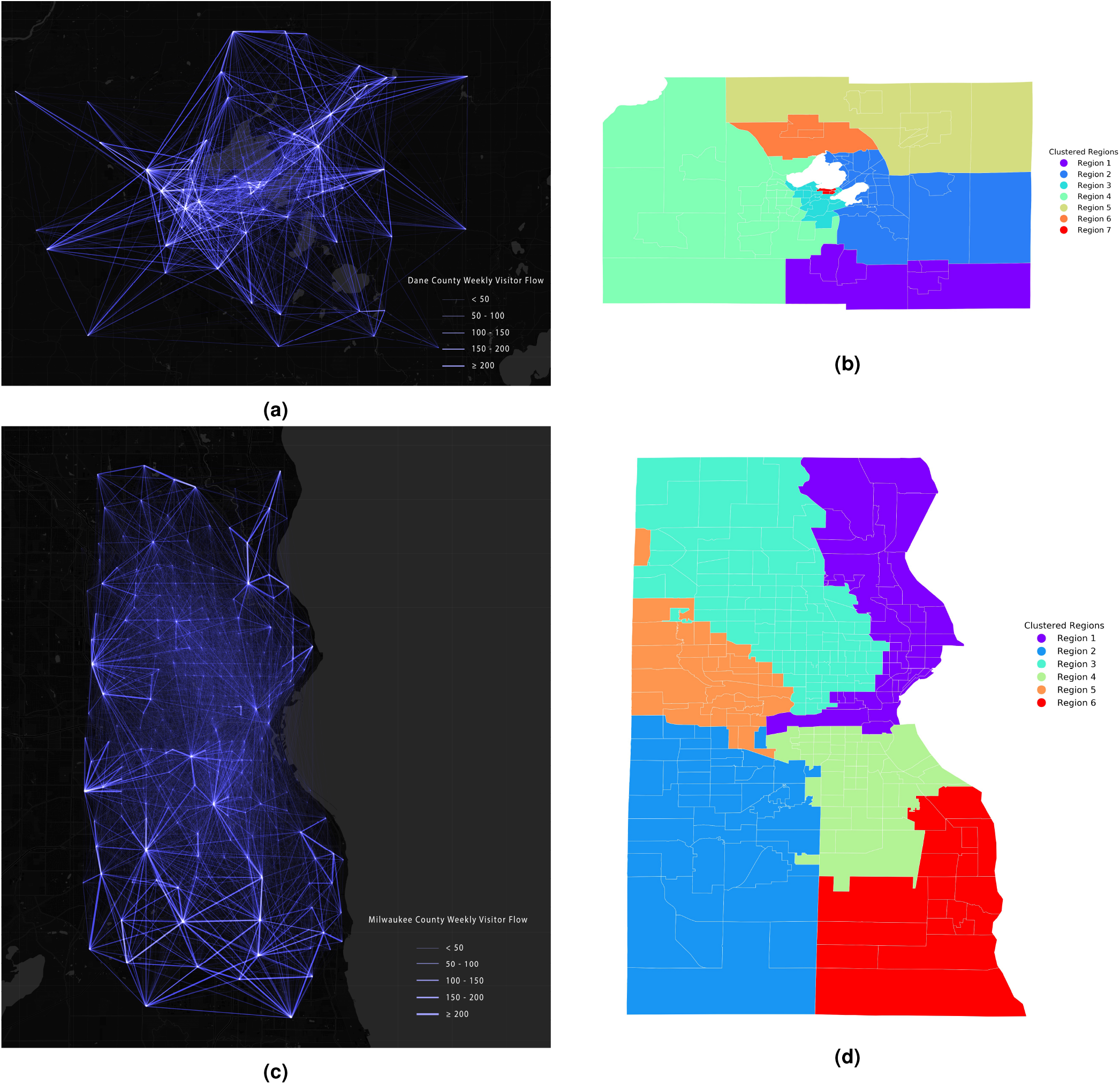
(a) Human movement O-D flows between census tracts in Dane County; (b) Dane county spatial clustering results using the Walktrap network community detection method; (c) Human movement O-D flows between census tracts in Milwaukee County; (d) Milwaukee County spatial clustering results using the Walktrap network community detection method.

### 4.2 Human mobility flow-augmented stochastic SEIR model results

#### Dane County

We treat the meta-population as relatively homogeneous within each region. We then build a separate SEIR model for each region, and couple these models together using the inter-regional mobility flow traffic information. Each region then has its own effective reproduction number *R*_*e*_, a function of time representing the expected number of new cases directly generated by each existing case. The effective reproduction number is a critical indicator that quantifies the spread of disease, and its dependence on time clearly is related to the county’s own “stay-at-home” order and subsequent reopening procedure. In Dane County, Phase 1 reopening was enacted on May 26. During this period, all business were allowed to open with 25% limit capacity. Phase 2 reopening was enacted on June 15, and the capacity was increased to 50%. On July 2, the county, after seeing an unusual increase of infection, rolled back the Phase 2 reopening and the capacity limit was reduced to 25%. On July 13, the county imposed an indoor face mask requirement.

In Figure 2, we plot the 3-day average *R*_*e*_ in each region and its corresponding interquartile range (IQR, the range that shows 25th to 75th percentiles). The dates of the reopening/rolling-back are also indicated in the plots. Region 7 (the downtown and university campus area) has the highest *R*_*e*_, spiking as high as 8 at the end of June, during the brief period of relaxed capacity limits for indoor businesses. By contrast, Region 3 (the adjoining residential area), despite also possessing a high population density, has a substantially lower *R*_*e*_, without a major late-June spike. In Figure 3a we plot the the traffic normalized effective reproduction number in Dane County. This measure controls for the fact that some regions have higher intra-regional traffic and communications than the others and are thus expected to have a higher *R*_*e*_. However, Region 7, even upon this normalization, still has a significantly higher *R*_*e*_ than all other regions, suggesting that there are social characteristics beyond population mobility contributing to rapid spread there. In particular, the normalized *R*_*e*_ has a peak in late June, approximately seven days after the county started Phase 2 reopening on June 15.

**Figure 2.**
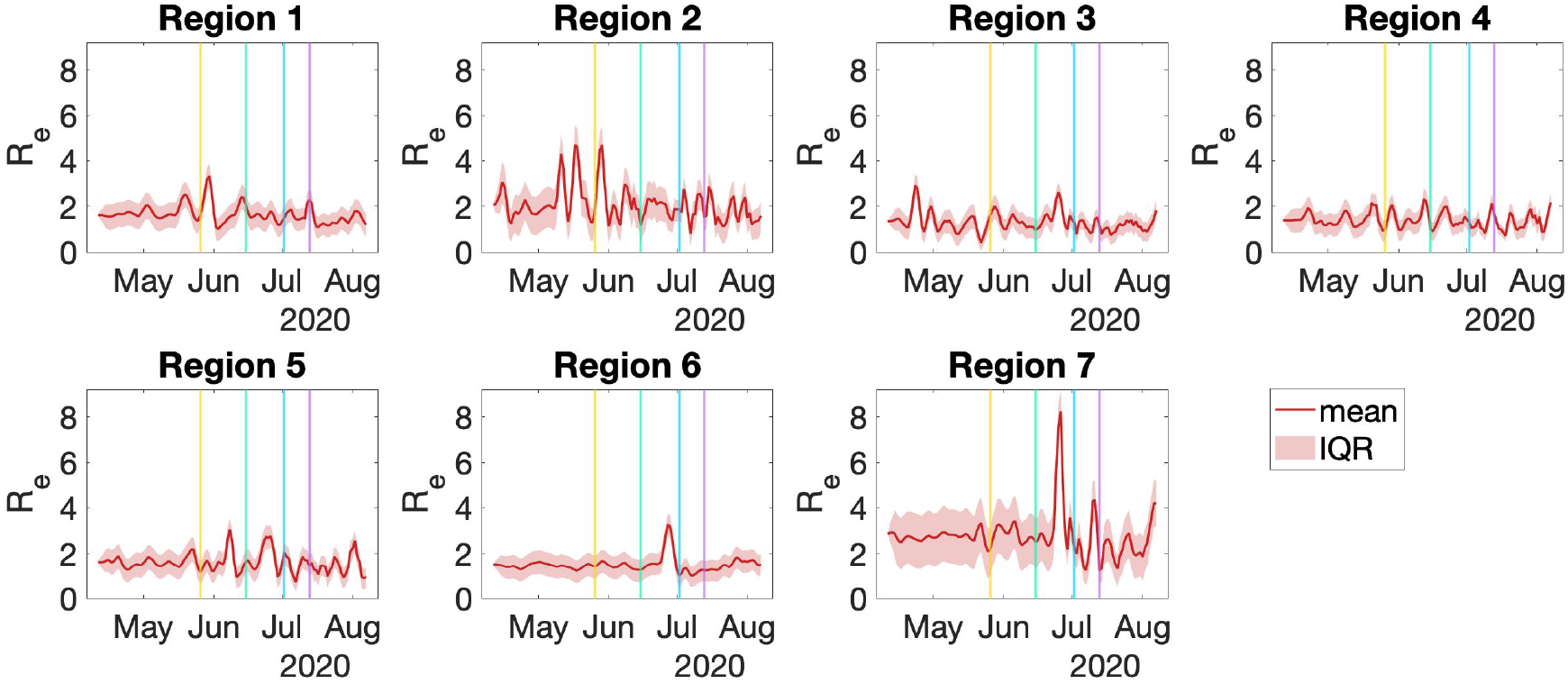
Effective reproduction number (3 day average) for different regions in Dane County. The vertical lines indicate the dates of Phase 1 reopening, Phase 2 reopening, rolling back reopening, and face covering order.

**Figure 3.**
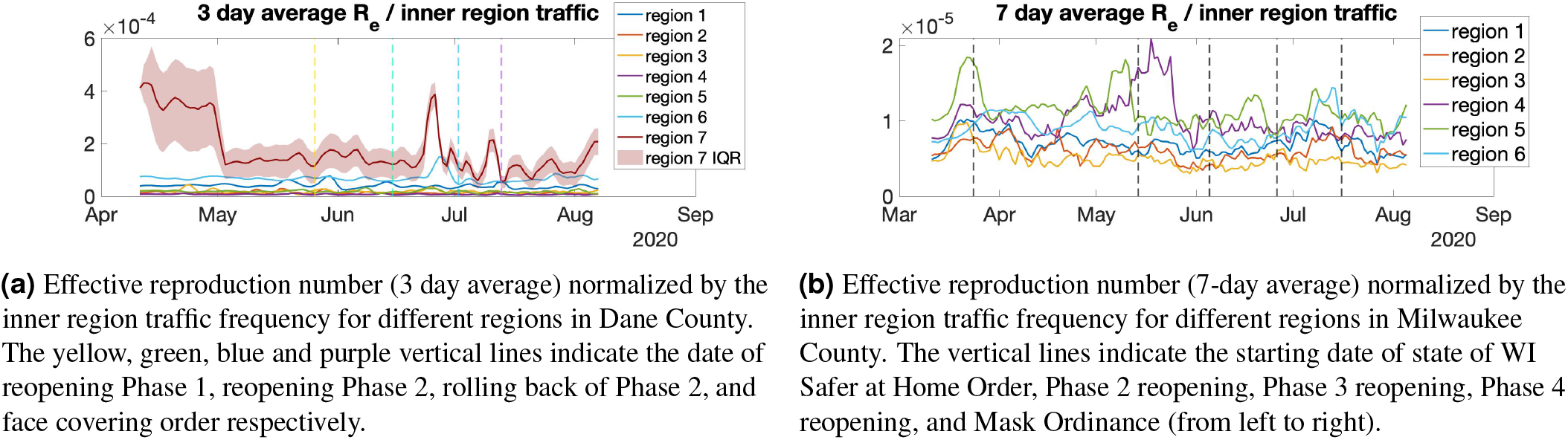
Effective reproduction number divided by inner region traffic.

The mobile device visit tracking data available to use allows to investigate some potential explanations for the differences of *R*_*e*_ between regions. In Table 3 we document the Pearson’s correlation of *R*_*e*_ in each region with different categories of business service visits. Clearly, the increase of *R*_*e*_ in Region 7 has a strong cross-correlation with the visits to “Drinking Places (Alcoholic Beverages)” with a time lag of 5 days. Such a data-driven analysis may complement the local official report showing that more than 21% of the newly COVID-19 infected cases reporting recent trips to bars and taverns^51^.

**Table 3.** The cross-correlation of effective reproduction number (3 day average) of different regions and the number of service visits (3 day average) within each region. The cross-correlations are computed using 5 days of time lag.

| Categories | Region 1 | Region 2 | Region 3 | Region 4 | Region 5 | Region 6 | Region 7 |
| --- | --- | --- | --- | --- | --- | --- | --- |
| Snack and Nonalcoholic Beverage Bars | -0.2520 | -0.1740 | -0.1850 | -0.1610 | -0.2250 | -0.1220 | -0.0830 |
| Drinking Places (Alcoholic Beverages) | -0.3410 | -0.1130 | 0.1180 | -0.1770 | 0.0140 | - | 0.3700 |
| Supermarkets and Grocery Stores | -0.0270 | 0.1130 | 0.4100 | 0.1610 | 0.1310 | -0.0840 | 0.1760 |
| Nature Parks and Other Similar Institutions | -0.0800 | 0.2100 | -0.1280 | 0.1720 | 0.1440 | 0.2900 | -0.0300 |
| Fitness and Recreational Sports Centers | -0.1990 | -0.3250 | 0.1520 | -0.0010 | 0.0850 | 0.0200 | 0.1370 |
| Full-Service Restaurants | -0.2780 | -0.2150 | 0.2540 | -0.0940 | 0.0820 | 0.0550 | 0.2950 |
| Limited-Service Restaurants | 0.0120 | -0.1580 | 0.1220 | -0.1290 | -0.1150 | -0.1020 | 0.2580 |

We also perform scenario studies to evaluate the possible consequences were different policies to be put in place. In particular, we study what would have happened if

- (case 1) there was no Phase 2 reopening enacted on June 15;
- (case 2) the county did not roll back the Phase 2 reopening on July 2;
- (case 3) the county further reopens on August 4 at different levels.

In Figure 4a we plot the predicted infection of selected regions supposing the June Phase 2 reopening had not taken place. Our model predicts clearly that the number of cases would be drastically reduced in Regions 3, 6, and 7 if the county did not enter Phase 2 reopening; in other words, those are the regions most severely affected by that decision. In particular, in Region 7, had there been no reopening, the projected number of cases is only 90 by June 30, as compared to the actual count of 282, more than three times as high. Regions 1, 2, 4, and 5 see small impacts from the reopening (see the results plotted in S9, S10 and S11.) In Figure 4b we plot the predicted infection assuming there was no rolling-back from Phase 2 reopening in the same three regions. It is clear that rolling-back the reopening reduced the number of infection cases significantly. Without rolling-back, the predicted total number of infection in Dane County during July 2-9 would have been 3, 394. This is almost three times of 1, 272, the actual number of confirmed cases in that time interval. In Figure 4c, we plot the predicted number of infections from August 4 into the future by 10 days. If the basic reproduction number tripled, the predicted number of infection would increase from 462 to 1,384 within 10 days.

**Figure 4.**
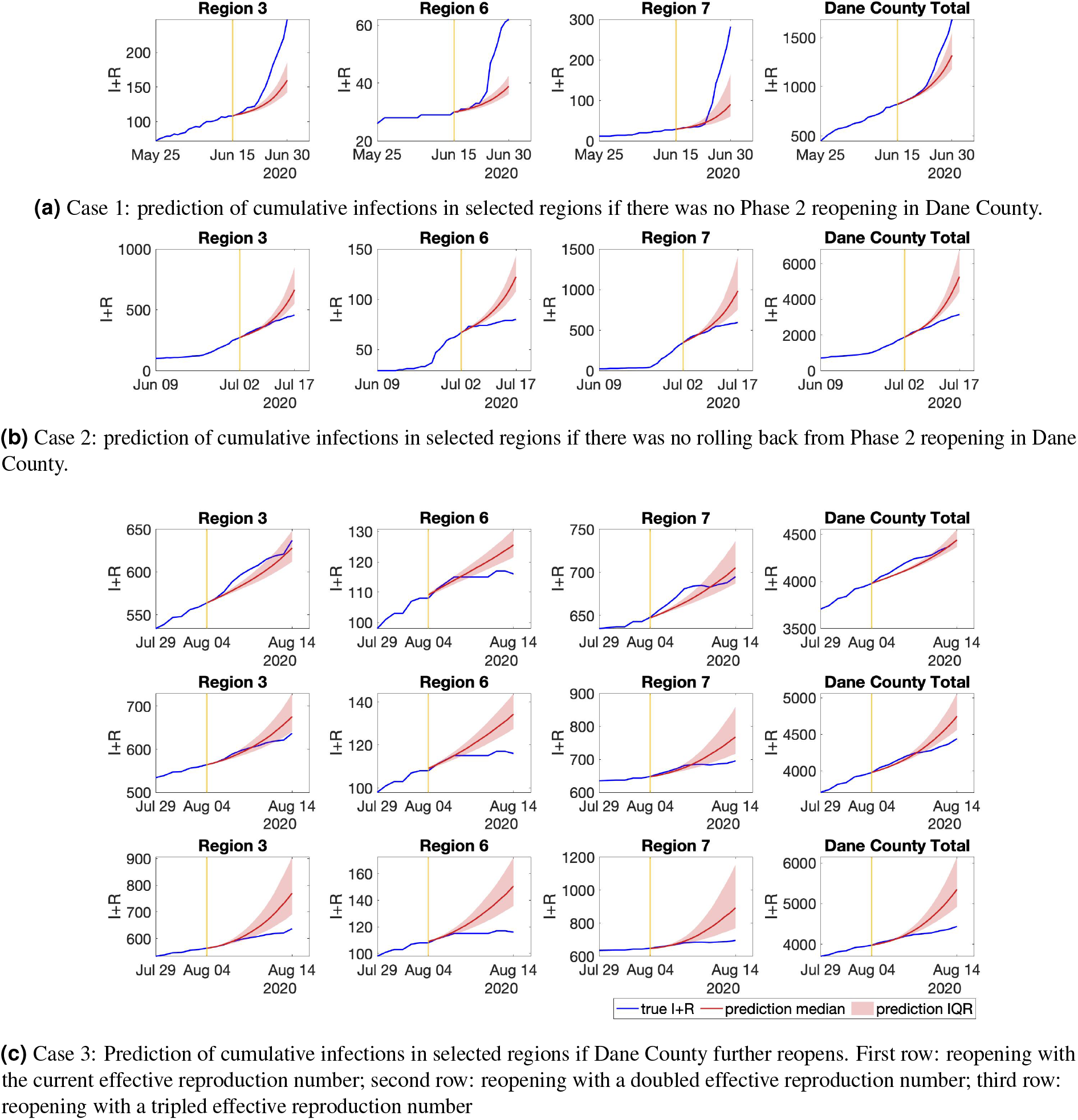
Prediction of cumulative infections if Dane County (a) did not enter Phase 2 reopening on June 15, (b) did not roll back from Phase 2 reopening on July 2 or, (c) further reopens on August 4.

To sum up: a strong cross-correlation is observed between the number of visits to “Drinking places (Alcoholic Beverages)” and increased infections in Region 7. At the same time, the scenario study suggests that without Phase 2 reopening, the infection in Dane county would have been significantly lower (reduce about two-thirds), and without the subsequent roll-back from Phase 2 reopening, the infection would have continued to increase dramatically. All this together provides strong evidence that the Phase 2 reopening allowing 50% capacity in drinking places played a major role in the drastic increase of the number of cases observed in Dane County during the summer of 2020.

#### Milwaukee County

We perform similar studies in Milwaukee County. The city of Mulwaukee started its Phase 2 reopening on May 14. During this period, restaurants and bars were allowed to have take-out or delivery service only. On June 5, Phase 3 reopening started, and restaurants and bars opened with 25% limit capacity. The limit capacity was lifted to 50% on June 26, when Phase 4 began. On July 16, Milwaukee instituted a mask ordinance requiring face covering in public space, both indoors and outdoors.

In Figure 5 we plot the effective reproduction number in different regions, and the corresponding IQR. Among the six regions, Regions 3 and 4 see the highest *R*_*e*_, averaging 1.8 and 2 respectively. In particular, in Region 4, in mid-May, *R*_*e*_ rose as high as 6. As mentioned above, Regions 3 and 4 are the two regions most subject to racial and ethnic segregation. Such neighborhood segregation patterns have also been identified previously through human mobility-based spatial interaction network analysis using location big data^52^. The COVID-19 spread and deaths in Milwaukee shows the racial disparities in health.

**Figure 5.**
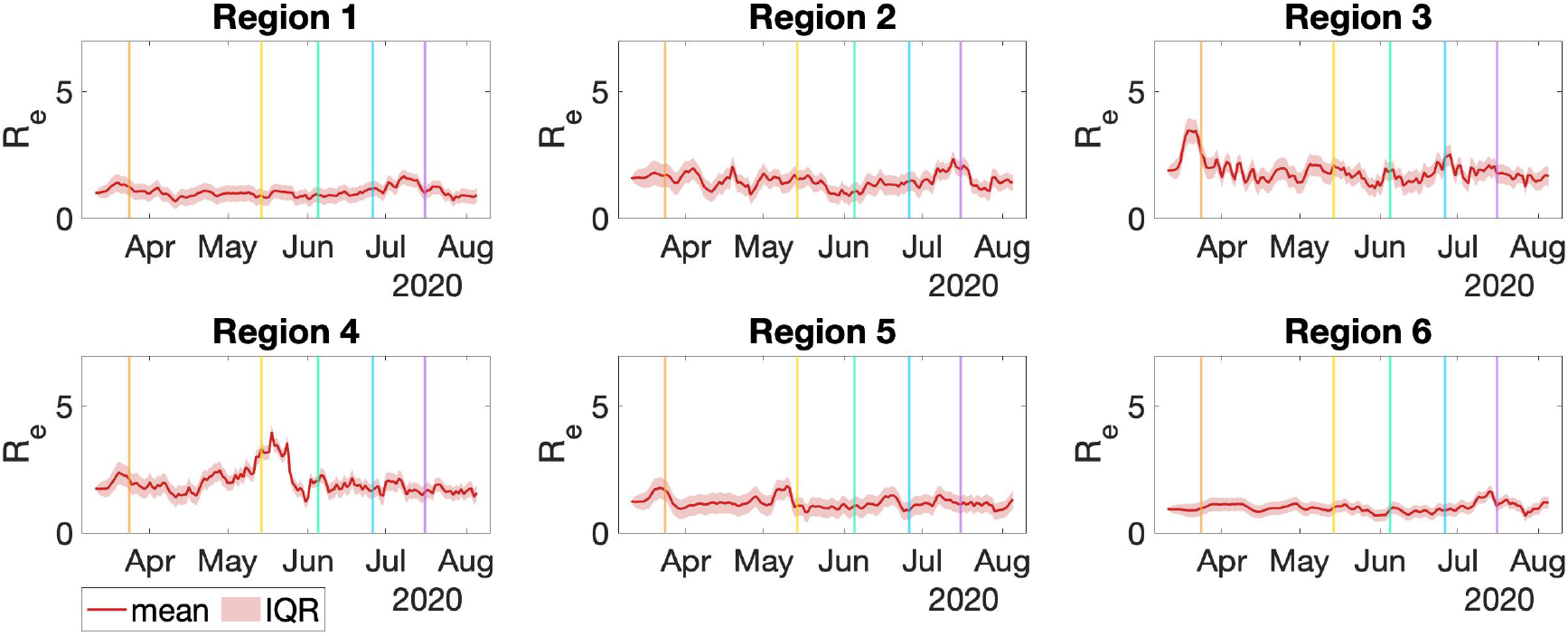
Effective reproductive number (7-day average) of Region 1 to Region 6 in Milwaukee County (left to right, top to bottom). The vertical lines indicate the starting date of state of WI Safer at Home Order, Phase 2 reopening, Phase 3 reopening, Phase 4 reopening, and Mask Ordinance (from left to right).

As we did in the Dane County study, we also consider the effective reproduction number normalized to control for intra-regional traffic. As seen in Figure 3b, even after this normalization, Region 4 still demonstrates the highest transmission. That is, even with high traffic frequency within this region being discounted, the local infection rate is still very high, with the mid-May peak persisting. This suggests that policymakers need to explicitly consider local transmission variation contexts even after restricting intra-regional traffic for preventing more health disparities in future pandemics.

### 4.3 Assessing Spatial Heterogeneity with Age, Race and People’s Self-Reported Social Distancing

The disparities in COVID-19 spread rate between different regions of the two counties studied reflect important interaction between demographic divisions and reaction to COVID-19. In Dane County, we observed an appreciable difference in the outcomes between people who lived near the campus area and those farther away, i.e., spatial heterogeneity. In Milwaukee County case, we observed stark differences in the burden of disease between different race & ethnicity groups, especially between predominantly white regions and regions with a higher Black and Hispanic population. One author in this manuscript was on the data analysis team of the “COVID-19 and Social Distancing” survey conducted between March 19th and April 1st, 2020 by a group of interdisciplinary scholars and non-governmental organizations in Wisconsin^53^. The survey received responses from over 30,000 Wisconsin residents. We conducted data analysis on a behavioral question asking people to what extent they practice social distancing and compared different age groups’ answers and different ethnic groups’ answers. We found that among respondents aged 20-29, a substantially higher proportion of participants (4.25%) said they did not perform social distancing at all or performed little social distancing, compared to the other two age groups (30-59: 1.26%, above 60: 1.03%) [Figure 6a]. For Hispanic and the African American ethnic groups, there is still certain amount of respondents who reported that they did not perform social distancing at all (the top red bar in Figure 6b). This self-reported behavioral differences in precautionary behavior might partially explain the heterogeneous patterns we observed in the two counties. More importantly, this disparities among age groups and race & ethnicity groups in the COVID-19 outcome and in the self-reported social distancing behavior flags an urgent policy need. Policymakers and health communicators must investigate the underlying reasons for demographically specific barriers to adoption of social distancing and other mitigation measures^54^.

**Figure 6.**
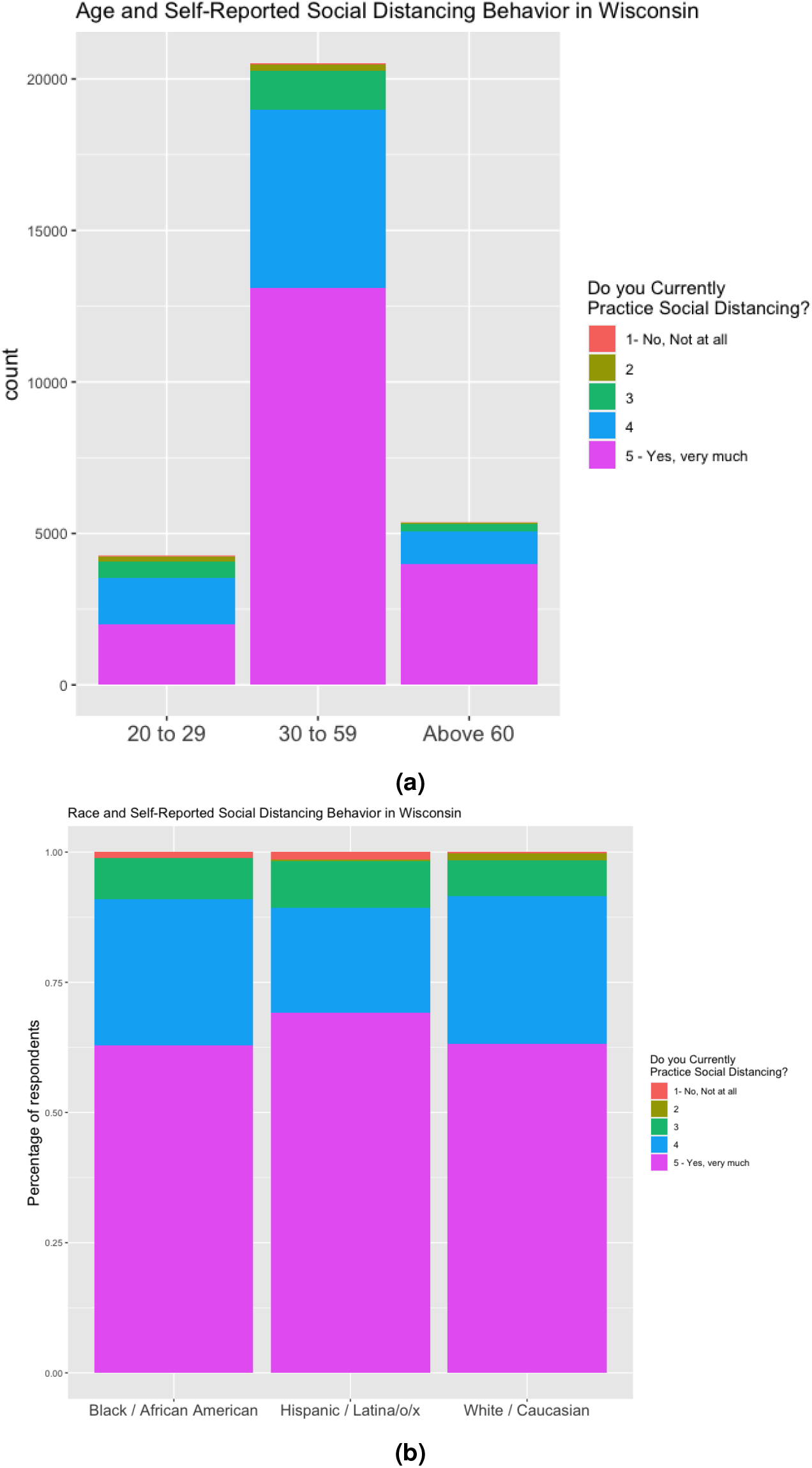
(a) Comparison on self-reported social distancing behavior among different age groups; (b) Comparison on self-reported social distancing behavior among different race & ethnicity groups.

## 5 Discussion

### 5.1 Sensitivity test of clustering

The results of clustering are subjected to the weekly travel O-D flow data and to the network-based community detection algorithms. In general, the clustering results in Milwaukee County and Dane County are rather stable with respect to the change of clustering methods and data from different weeks. We test the sensitivity of clustering using different clustering methods and different periods of data.

The results shown above utilize the Walktrap method on the travel O-D flow data of the week of March 2. The Walktrap algorithm uses short random walks to detect communities in a large graph. In comparison, we also apply the Louvain method on the same data. The Louvain method is a popular community detection algorithm that aims to maximize the modularity^55^. Figures S2 and S3 show the clustering results of Milwaukee County and Dane County using the Louvain method. For Milwaukee County, the two clustering results are very similar apart for a small section in the lower-left corner of the map. Both methods result in 6 clustering groups. The modularities of clustering using Walktrap and Louvain are 0.365 and 0.376 respectively, indicating almost identical performance. For Dane County, however, the two methods provide slightly different results. Walktrap divides the county into 7 areas, while the Louvain gives 5. Comparing the subfigures in Figure S3, Louvain method clusters the central area as a single region, but the Walktrap method further divides this area into smaller components. Region 7 is the downtown area where the campus of University of Wisconsin – Madison is located. It plays an important role in our analysis of human mobility flows and infection of COVID-19, and thus it is meaningful to identify it as a single region. We therefore chose to use the Walktrap method. The modularities of clustering using Walktrap and Louvain algorithms are very close: 0.318 and 0.334 respectively.

In addition to comparing different clustering methods, we also compare the clustering results using data from different time intervals. In Figures S2 and S3 we show the results for the clustering of two counties using the data of March 2-8, and 9-15 respectively. For Dane County, only 3 out of 105 census tracts (each small sub-region in the figure represents a census tract) appear in different groups after the change of data. In Milwaukee County, after changing the data, 13 out of 296 census tracts have different group assignment using Louvain method 17 of 296 census tracts were assigned to a different cluster using Walktrap method. This indicates the clustering results are stable enough for the purpose of this project.

### 5.2 Policy implications

Our results suggest several policy lessons. Consider Dane County, which contains a large “college town” environment. As we see in Figure 3a, the vertical dashed line which corresponds to the policy phases of re-opening in Dane County, the area experienced spikes in the reproduction number when the county entered the Phase 2 reopening in mid June. We also observed that the reproduction number decreased significantly after the county rolled back its reopening policy in early July. The pattern of reproduction rate following the policies is especially substantial for the campus region. These results suggest that policymakers need to design reopening in a more nuanced manner. Instead of implementing an one-size-fits-all reopening policy for all the regions within a county, reopening policies that attend to the diverse nature of regions within a county should be considered. Policymaking needs to be spatially heterogeneous. For instance, we demonstrated that certain types of business and certain spatially distinct regions have a much higher infection rate in Dane County: visits to drinking places have the strongest association with infection rates, and the campus region has a higher infection rate than other parts of the county do. These empirical evidences flag the crucial importance of designing reopening policies which take these particularities into account when choosing what types of business and what regions to reopen first. Public policies also need to adapt to different regions that consist of very different demographic populations in order to control the reproduction rate of COVID-19, since COVID-19 is not only a public health problem, but also a problem that reflects existing disparities among various socially distinct population groups. For instance, for testing supply and future vaccination enforcement, policymakers might prioritize target certain demographic regions with priority.

## Data Availability

The official testing results of COVID-19 confirmed cases between March 11, 2020 and August 12, 2020 were obtained from the local COVID-19 Dashboards, created by the Public Health Offices of City of Madison & Dane County (https://publichealthmdc.com/coronavirus) and Milwaukee County (https://county.milwaukee.gov/EN/COVID-19). The census-tract level geographic boundaries with demographics and socioeconomic attributes were obtained from the U.S. Census Bureau. We collected points of interest (POIs) with aggregated foot-traffic information from SafeGraph. For each POI, the records of aggregated visitor patterns record the number of unique visitors and the number of total visits to each venue during a specified time window (i.e., hourly, weekly, and monthly); this allows us to estimate the foot-traffic of each venue and the origin-to-destination (O-D) spatial interaction flow patterns during the the study period (https://github.com/GeoDS/COVID19USFlows). We further aggregate the O-D flow matrices to the census-tract level to match the COVID-19 testing data.

https://county.milwaukee.gov/EN/COVID-19

## Acknowledgements

We would like to thank the SafeGraph Inc. for providing the anonymous and aggregated place visits and human mobility flow data. S.G., Q.L., K.C, and J.P. acknowledge the funding support provided by the National Science Foundation (Award No. BCS-2027375). J.E. acknowledge the funding support provided by the National Science Foundation (Award No. DMS-1700884). Q.L., N.C. and X.H. are also supported by Data Science Initiative, provided by the University of Wisconsin - Madison Office of the Chancellor and the Vice Chancellor for Research and Graduate Education with funding from the Wisconsin Alumni Research Foundation. Any opinions, findings, and conclusions or recommendations expressed in this material are those of the author(s) and do not necessarily reflect the views of the National Science Foundation.

## Author contributions statement

Research design and conceptualization: S.G., Q.L., N.C; Data collection and processing: X.H., J.R., Y.K.; Result analysis: X.H, S.G., Q.L., N.C, K.C., J.E.; Visualization: H.X, J.R., Y.K., K.C.; Obtained funding: S.G.,Q.L.,N.C,J.P.,J.E.; Project administration: S.G., J.P.; Writing: all authors.

## Footnotes

1 www.safegraph.com/blog/what-about-bias-in-the-safegraph-dataset

## References

1. (2020). Centers for Disease Control and Prevention, COVID-19 Cases in U.S. Centers for Disease Control and Prevention. Accessed August 24, 2020, https://www.cdc.gov/coronavirus/2019-ncov/cases-updates/cases-in-us.html.

2. Pan, A. et al.. Association of public health interventions with the epidemiology of the COVID-19 outbreak in wuhan, china. JAMA Online First (2020).

3. Hartley, D. M. & Perencevich, E. N. Public health interventions for COVID-19: Emerging evidence and implications for an evolving public health crisis. JAMA 0098–7484 (2020).

4. Lai, S. et al.. Effect of non-pharmaceutical interventions to contain COVID-19 in china. Nature (2020).

5. Chinazzi, M. et al.. The effect of travel restrictions on the spread of the 2019 novel coronavirus (COVID-19) outbreak. Science (2020).

6. Tian, H. et al.. An investigation of transmission control measures during the first 50 days of the covid-19 epidemic in china. Science (2020).

7. Zhang, R., Li, Y., Zhang, A. L., Wang, Y. & Molina, M. J. Identifying airborne transmission as the dominant route for the spread of COVID-19. Proc. Natl. Acad. Sci. (2020).

8. Duque, D. et al.. Timing social distancing to avert unmanageable covid-19 hospital surges. Proc. Natl. Acad. Sci. 117, 19873–19878 (2020).

9. Giordano, G. et al.. Modelling the COVID-19 epidemic and implementation of population-wide interventions in italy. Nat. Medicine 1–6 (2020).

10. Gatto, M. et al.. Spread and dynamics of the COVID-19 epidemic in italy: Effects of emergency containment measures. Proc. Natl. Acad. Sci. 117, 10484–10491 (2020).

11. Team, I. C.-.F. & Hay, S. Covid-19 scenarios for the united states. MedRxiv (2020).

12. Team, I. C.-. F., Murray, C. J. et al.. Forecasting COVID-19 impact on hospital bed-days, ICU-days, ventilator-days and deaths by us state in the next 4 months. MedRxiv (2020).

13. Zhou, Y. et al.. Effects of human mobility restrictions on the spread of COVID-19 in Shenzhen, China: a modelling study using mobile phone data. The Lancet Digit. Heal. 2, e417–e424 (2020).

14. Aleta, A. et al.. Modelling the impact of testing, contact tracing and household quarantine on second waves of COVID-19. Nat. Hum. Behav. 1–8 (2020).

15. Alagoz, O., Sethi, A., Patterson, B., Churpek, M. & Safdar, N. Impact of timing of and adherence to social distancing measures on COVID-19 burden in the us: A simulation modeling approach. medRxiv (2020).

16. Kraemer, M. U. et al.. The effect of human mobility and control measures on the COVID-19 epidemic in China. Science (2020).

17. Fang, H., Wang, L. & Yang, Y. Human mobility restrictions and the spread of the novel coronavirus (2019-ncov) in China. Tech. Rep., National Bureau of Economic Research (2020).

18. Yan, L. et al.. An interpretable mortality prediction model for COVID-19 patients. Nat. Mach. Intell. 1–6 (2020).

19. Wang, P., Zheng, X., Li, J. & Zhu, B. Prediction of epidemic trends in COVID-19 with logistic model and machine learning technics. Chaos, Solitons & Fractals 110058 (2020).

20. Zhou, Y. et al.. A spatiotemporal epidemiological prediction model to inform county-level covid-19 risk in the united states. Harv. Data Sci. Rev. (2020).

21. Thomas, L. J. et al.. Spatial heterogeneity can lead to substantial local variations in COVID-19 timing and severity. Proc. Natl. Acad. Sci. pnas.2011656117 (2020).

22. Scheufele, D. A., Krause, N. M., Freiling, I. & Brossard, D. How not to lose the COVID-19 communication war. Issues Sci. Technol. 17 (2020).

23. Flynn, J., Slovic, P. & Mertz, C. K. Gender, race, and perception of environmental health risks. Risk analysis 14, 1101–1108 (1994).

24. Berrigan, D., Dodd, K., Troiano, R. P., Krebs-Smith, S. M. & Barbash, R. B. Patterns of health behavior in us adults. Prev. medicine 36, 615–623 (2003).

25. Scheufele, D. A. Science communication as political communication. Proc. Natl. Acad. Sci. 111, 13585–13592 (2014).

26. Baum, N. M., Jacobson, P. D. & Goold, S. D. “listen to the people”: public deliberation about social distancing measures in a pandemic. The Am. J. Bioeth. 9, 4–14 (2009).

27. Andersen, M. Early evidence on social distancing in response to COVID-19 in the united states. Available at SSRN 3569368 (2020).

28. Farber, S. & Johnson, J. New data shows young people need to take social distancing seriously. ABC News (2020).

29. Clark, W. A. Residential segregation in american cities: A review and interpretation. Popul. research Policy review 5, 95–127 (1986).

30. Chevan, A. Age, housing choice, and neighborhood age structure. Am. J. Sociol. 87, 1133–1149 (1982).

31. Rothstein, R. The color of law: A forgotten history of how our government segregated America (Liveright Publishing, 2017).

32. WashingtonPost (2020). “Income emerges as a major predictor of coronavirus infections, along with race, available at https://www.washingtonpost.com/health/income-emerges-as-a-major-predictor-of-coronavirus-infections-along-with-race/2020/06/22/9276f31e-b4a3-11ea-a510-55bf26485c93_story.html. Accessed on August 10, 2020.”.

33. Rubin, D. et al.. Association of social distancing, population density, and temperature with the instantaneous reproduction number of SARS-CoV-2 in counties across the united states. JAMA Netw. Open 3, e2016099–e2016099 (2020).

34. Van Bavel, J. J. et al.. Using social and behavioural science to support COVID-19 pandemic response. Nat. Hum. Behav. 1–12 (2020).

35. Weill, J. A., Stigler, M., Deschenes, O. & Springborn, M. R. Social distancing responses to COVID-19 emergency declarations strongly differentiated by income. Proc. Natl. Acad. Sci. 117, 19658–19660 (2020).

36. Barrios, J. M. & Hochberg, Y. V. Risk perception through the lens of politics in the time of the COVID-19 pandemic. Tech. Rep., University of Chicago (2020).

37. Allcott, H. et al.. Polarization and public health: Partisan differences in social distancing during the coronavirus pandemic. NBER Work. Pap. (2020).

38. (2020). The Brookings Institution, Black-white segregation in U.S. metro areas. Accessed September 27, 2020, https://www.brookings.edu/blog/the-avenue/2018/12/17/black-white-segregation-edges-downward-since-2000-census-shows/.

39. (2020). Robert Wood Johnson Foundation and University of Wisconsin Population Health Institute, The 2020 County Health Rankings, Accessed September 27, 2020, https://www.countyhealthrankings.org/explore-health-rankings.

40. (2020). Public Health Office of Dane County, Dane County of Wisconsin COVID-19 Dashboard. Accessed August 12, 2020, https://publichealthmdc.com/coronavirus.

41. (2020). Public Health Office of Milwaukee County, Milwaukee County of Wisconsin COVID-19 Dashboard. Accessed August 12, 2020, https://county.milwaukee.gov/EN/COVID-19.

42. Bureau, U. C. (2020). Cartographic Boundary Files - Shapefile. Accessed August 12, 2020, https://www.census.gov/geographies/mapping-files/time-series/geo/carto-boundary-file.html.

43. Survey, U. C. B. A. C. (2020). Cartographic Boundary Files - Shapefile. Accessed August 12, 2020, https://www.census.gov/programs-surveys/acs.

44. Kang, Y. et al.. Multiscale dynamic human mobility flow dataset in the us during the covid-19 epidemic. arXiv preprint arXiv:2008.12238 (2020).

45. Kucharski, A. J. et al.. Early dynamics of transmission and control of COVID-19: a mathematical modelling study. The Lancet Infect. Dis. (2020).

46. Pons, P. & Latapy, M. Computing communities in large networks using random walks. In International Symposium on Computer and Information Sciences, 284–293 (Springer, 2005).

47. Stuart, A. & Zygalakis, K. Data assimilation: A mathematical introduction. Tech. Rep., Oak Ridge National Lab.(ORNL), Oak Ridge, TN (United States) (2015).

48. Evensen, G. The ensemble kalman filter for combined state and parameter estimation. IEEE Control. Syst. Mag. 29, 83–104 (2009).

49. Reich, S. & Cotter, C. Probabilistic forecasting and Bayesian data assimilation (Cambridge University Press, 2015).

50. Moon, T. K. The expectation-maximization algorithm. IEEE Signal Process. Mag. 13(6), 47–60 (1996).

51. Rickert, C. & Wahlberg, D. Amid rise in covid-19 cases, dane county tightens restrictions on bars, restaurants, indoor gatherings. Wis. State J. (2020).

52. Prestby, T., App, J., Kang, Y. & Gao, S. Understanding neighborhood isolation through spatial interaction network analysis using location big data. Environ. Plan. A: Econ. Space.

53. Wirz, C., Schwakopf, J., Brossard, D., DiPrete Brown, L. & Brauer, M. Self-reported compliance and attitudes about social distancing during the COVID-19 outbreak. OSF Prepr. (2020).

54. Chen, K. et al.. How public perceptions of social distancing evolved over a critical time period: communication lessons learnt from the american state of Wisconsin. J. Sci. Commun. (forthcoming).

55. Blondel, V. D., Guillaume, J.-L., Lambiotte, R. & Lefebvre, E. Fast unfolding of communities in large networks. J. Stat. Mech. Theory Exp. 2008, P10008 (2008).

