## Supporting Information for "Intra-county modeling of COVID-19 infection with human mobility: assessing spatial heterogeneity with business traffic, age and race"

September 30, 2020

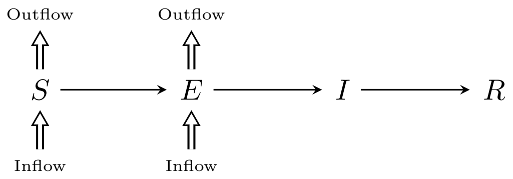


Figure S1: Flowchart of the human mobility flow-augmented stochastic SEIR model.

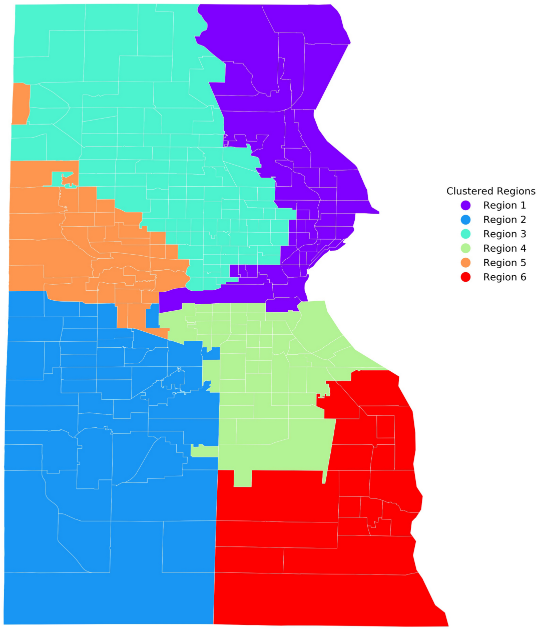


(a) Walktrap method, week of March 2, modularity 0.365

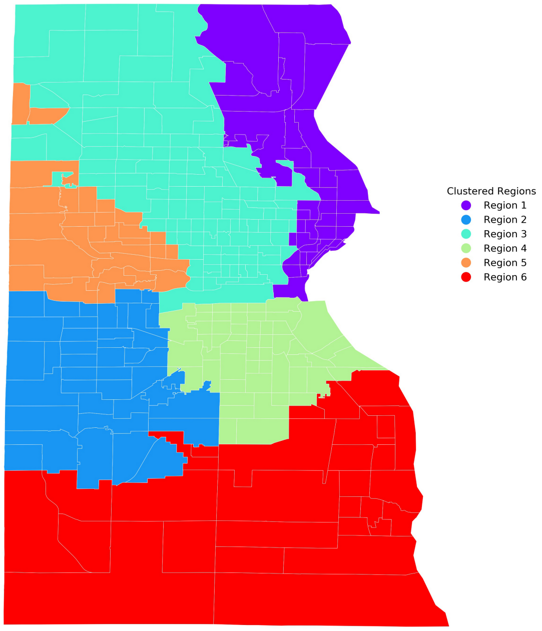


(b) Louvain method, week of March 2, modularity 0.376

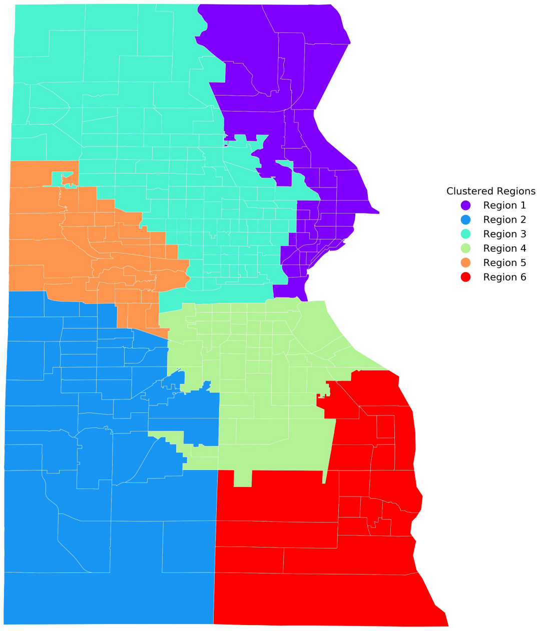


(c) Walktrap method, week of March 9, modularity 0.375

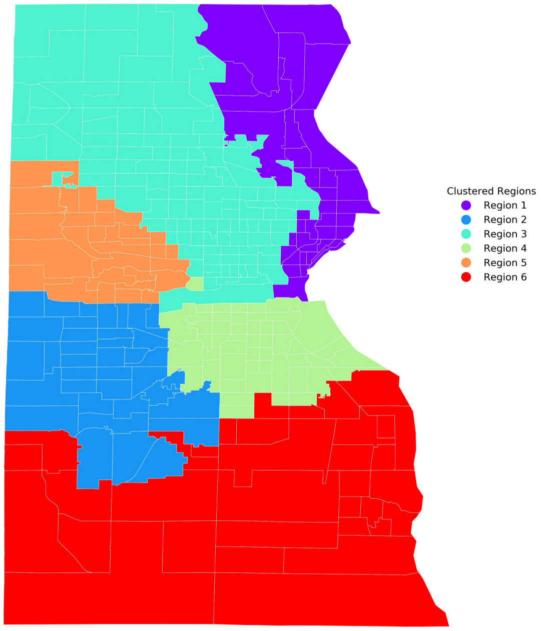


(d) Louvain method, week of March 9, modularity 0.378

Figure S2: Clustering results of Milwaukee County

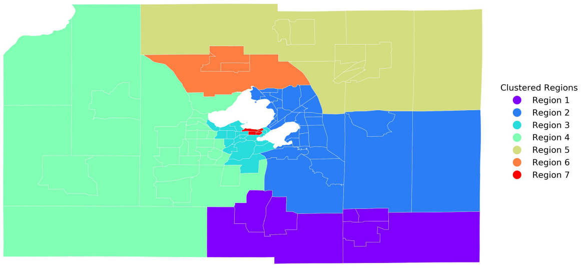


(a) Walktrap method, week of March 2, modularity 0.318

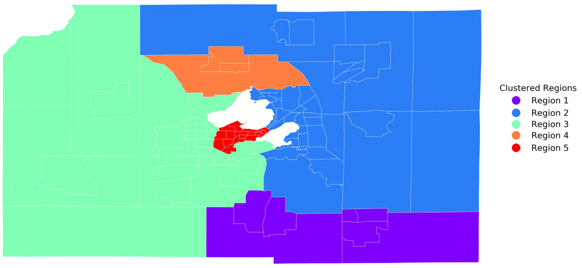


(b) Louvain method, week of March 2, modularity 0.334

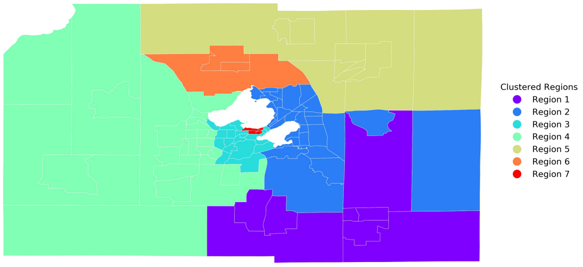


(c) Walktrap method, week of March 9, modularity 0.324

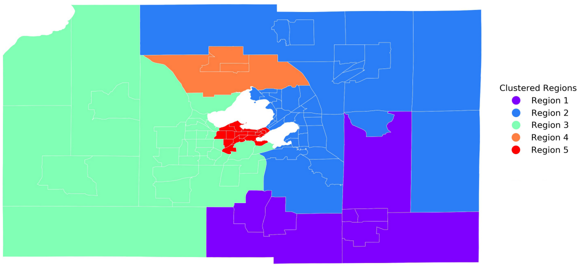


(d) Louvain method, week of March 9, modularity 0.343

Figure S3: Clustering results of Dane county

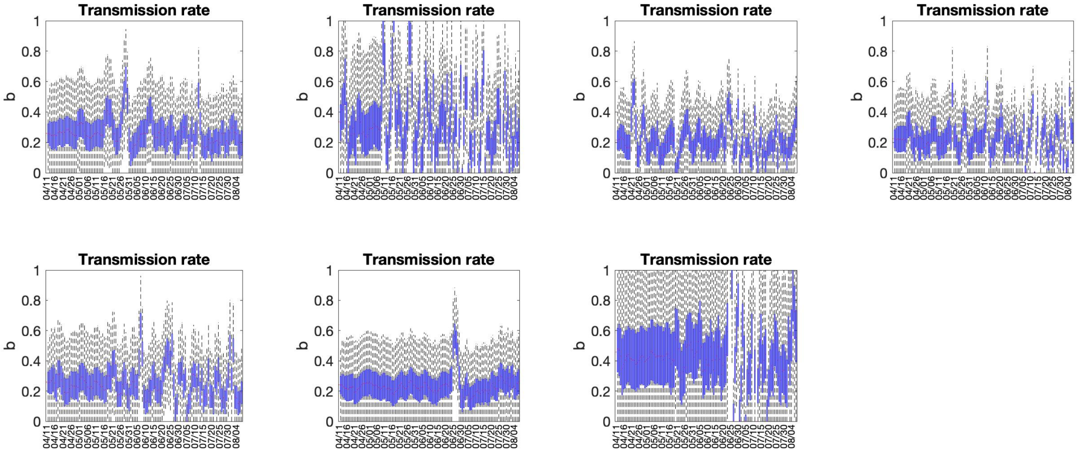


Figure S4: Transmission rate of different regions in Dane County.

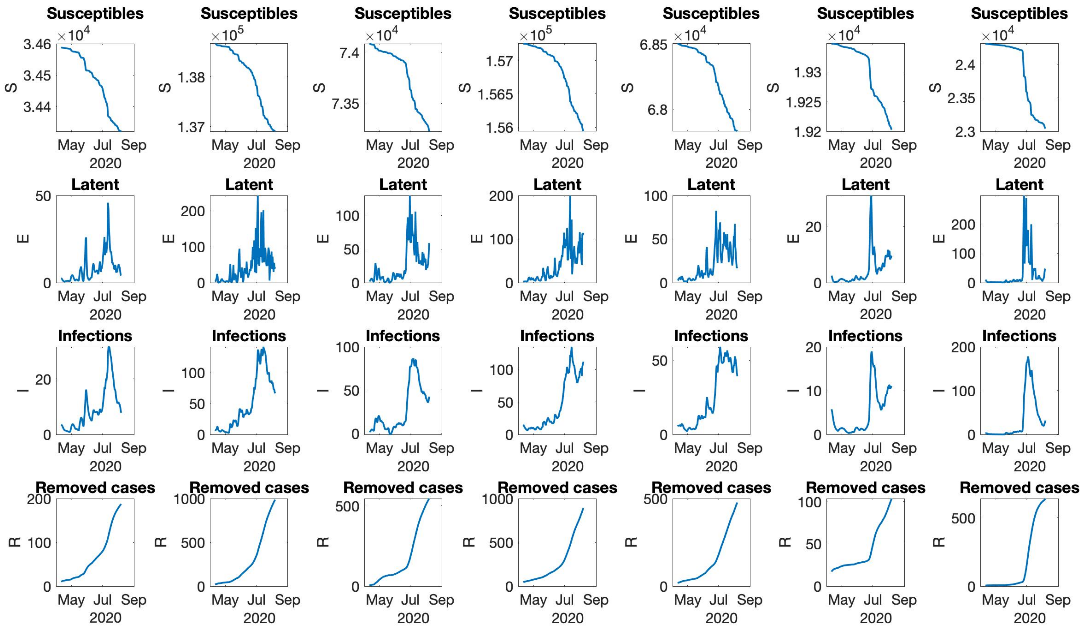


Figure S5: The evolution of $S$, $E$, $I$, and $R$ for Region 1 to Region 7 in Dane County (left to right, top to bottom).

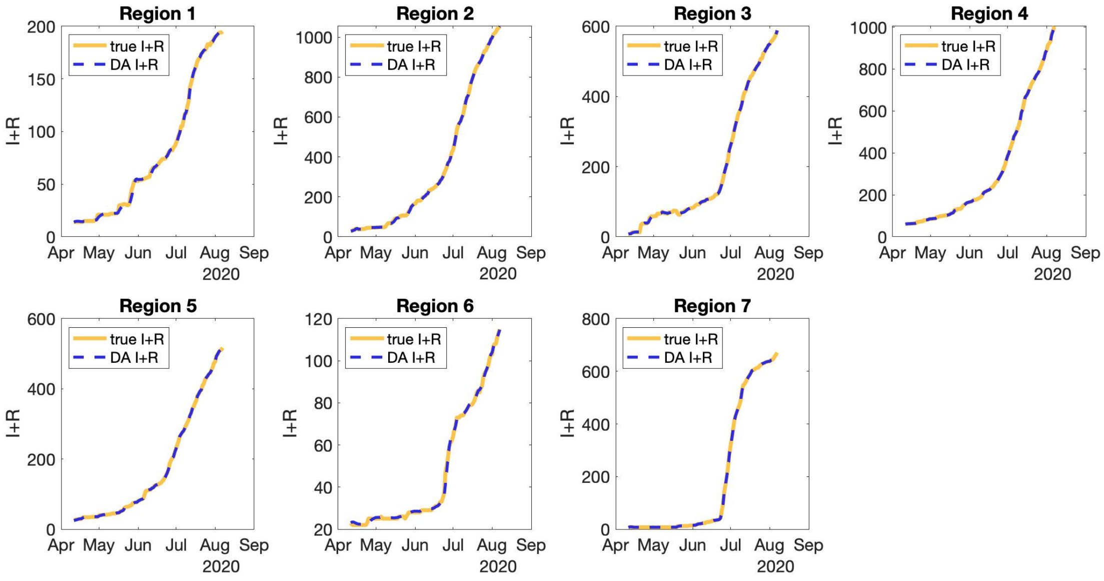


Figure S6: Data assimilation result and true cumulative infection data for Region 1 to Region 7 in Dane County.

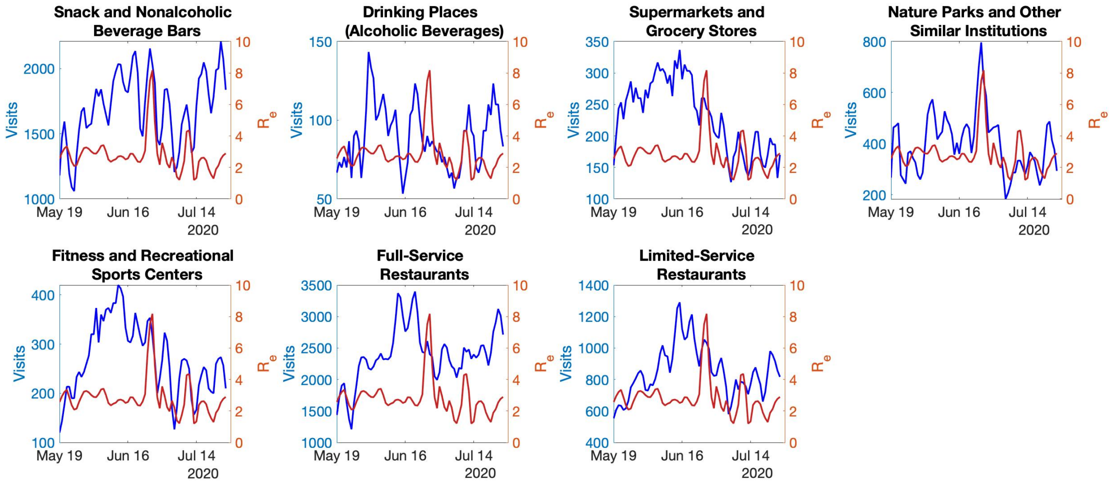


Figure S7: Number of visits (3 day average) to different categories of businesses in Region 7 and effective reproduction number (3 day average) of Region 7 from May 19 to July 26.

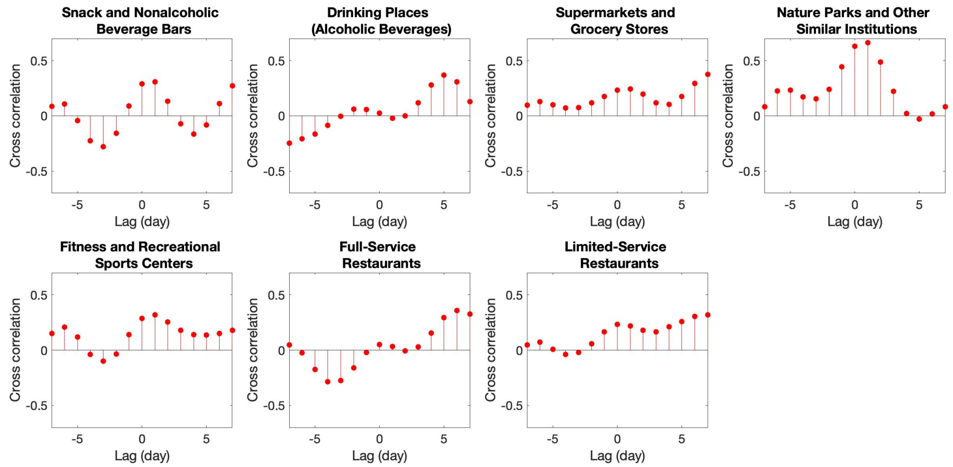


Figure S8: The cross-correlation of effective reproduction number (3 day average) of downtown Madison area (Region 7) and number of visits (3 day average) to different categories of businesses in Region 7 from May 19 to July 26. The cross-correlations are plotted based on different time lags.

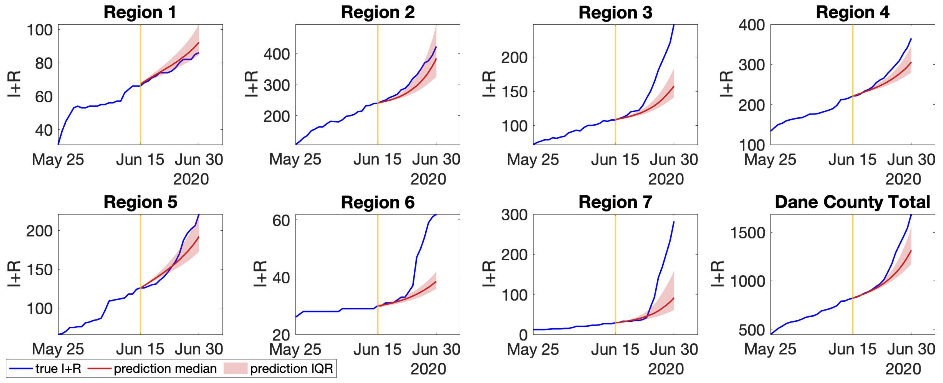


Figure S9: Case 1: prediction of cumulative infections in each region if there was no Phase 2 reopening in Dane County.

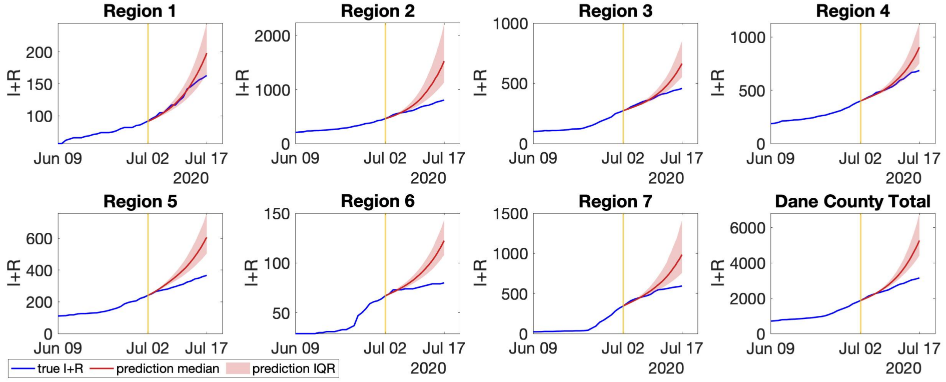


Figure S10: Case 2: prediction of cumulative infections in selected regions if there was no rolling back from Phase 2 reopening in Dane County.

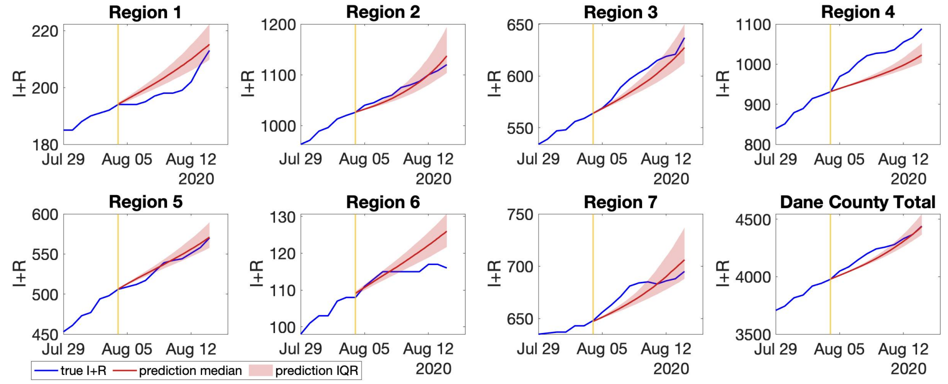


(a) Original effective reproduction number

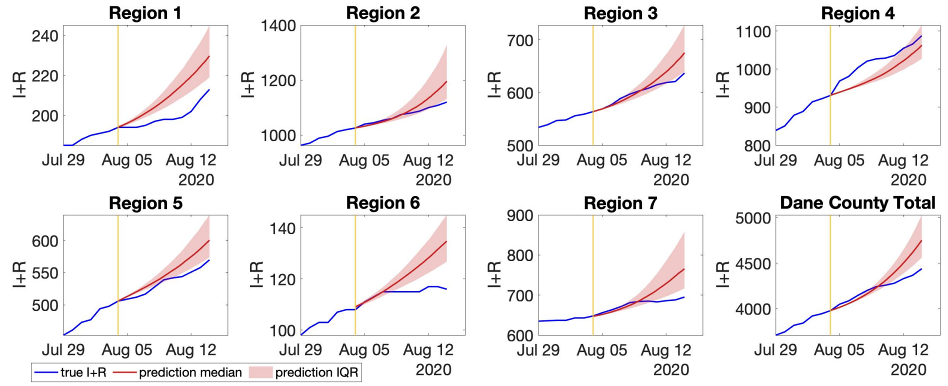


(b) Double effective reproduction number

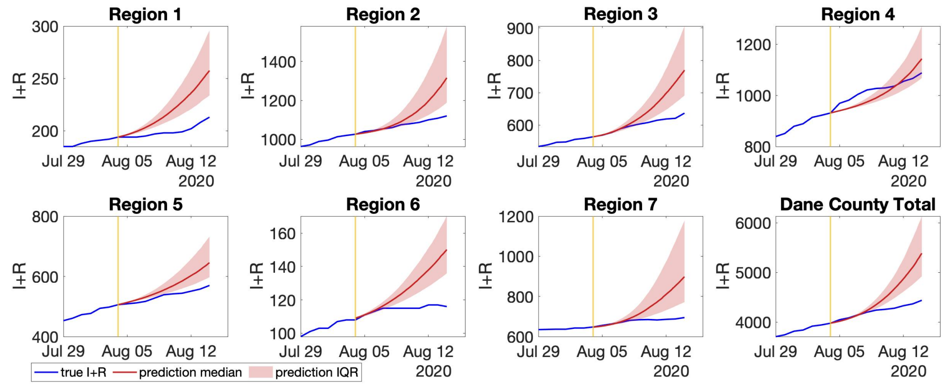


(c) Double effective reproduction number

Figure S11: Prediction of cumulative infections if Dane County further reopens.

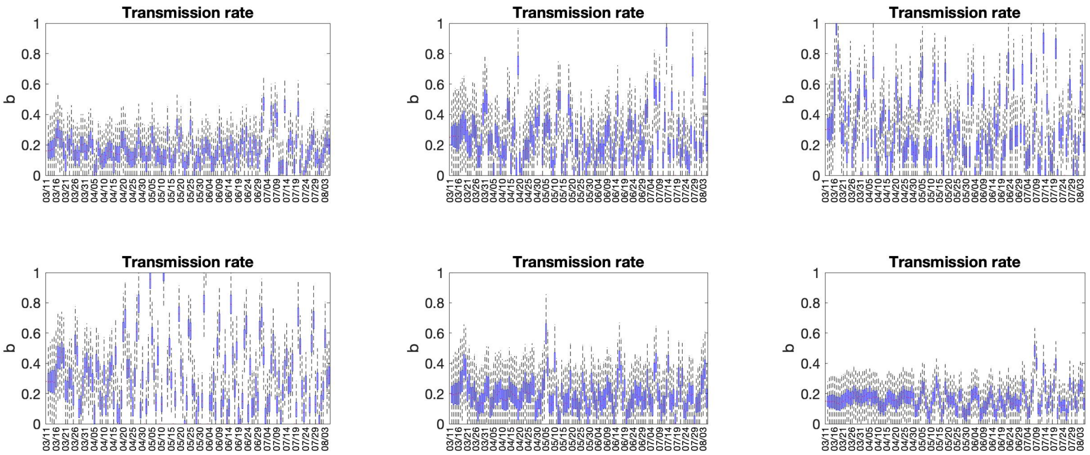


Figure S12: Transmission rate of different regions in Milwaukee County.

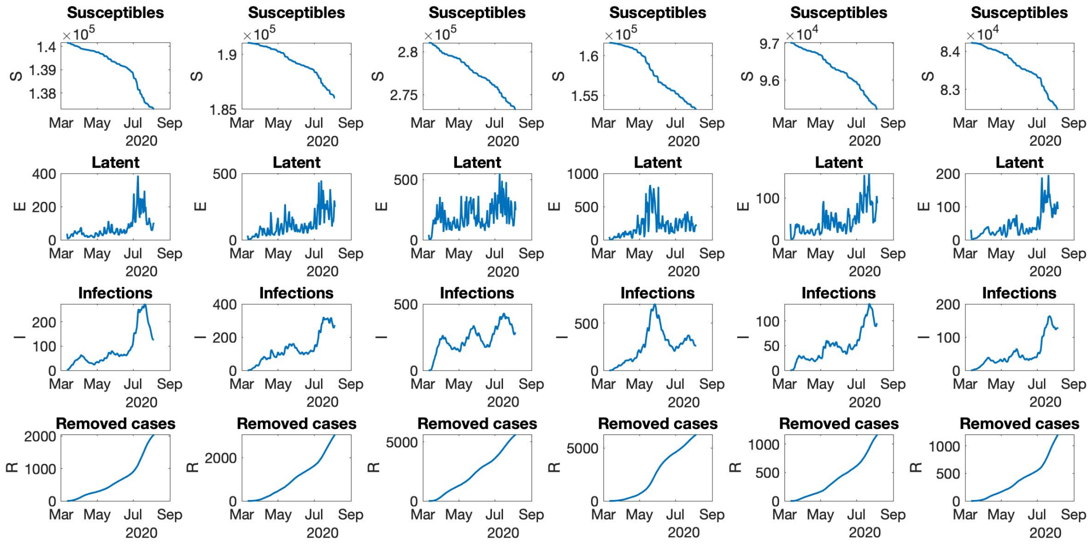


Figure S13: The evolution of $S$, $E$, $I$, and $R$ for Region 1 to Region 6 in Milwaukee County (left to right, top to bottom).

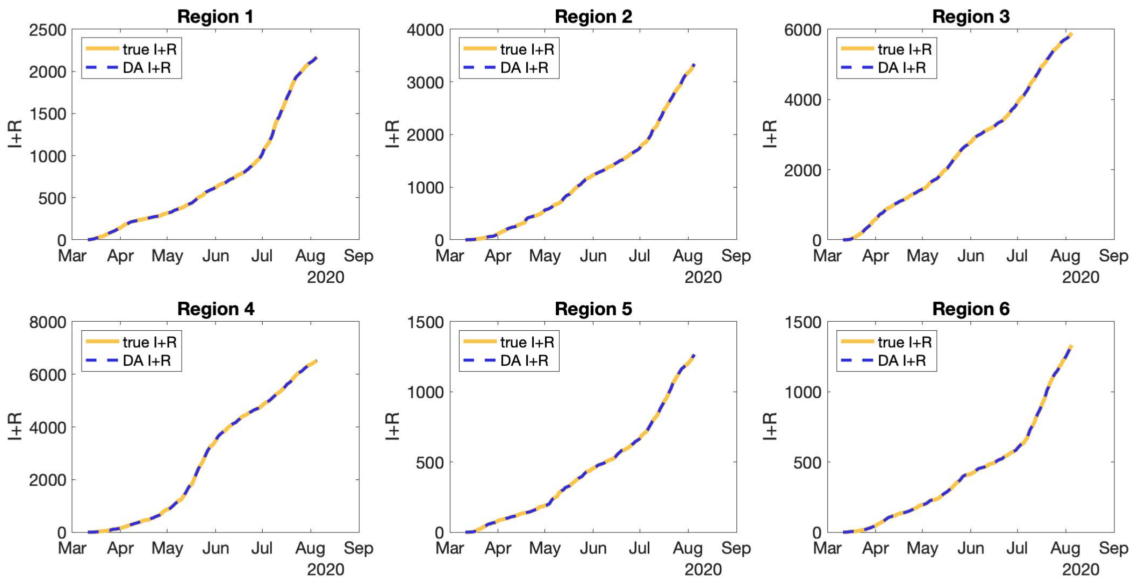


Figure S14: Data assimilation result and true cumulative infection data for Region 1 to Region 6 in Milwaukee County.
